# Early microvascular and neuro-retinal alterations in type 2 diabetic patients without diabetic retinopathy

**DOI:** 10.1101/2020.11.22.20236141

**Authors:** Wei Wang, Yingfeng Zheng, Sen Liu, Yuting Li, Wenyong Huang, Xiaolin Liang, Yizhi Liu

## Abstract

**Purpose:** to evaluate and correlate the alterations of microvascular and chorioretinal structure in Type 2 diabetes mellitus (T2DM) patients without clinical signs of DR.

**Methods:** T2DM patients were randomized sampled from Guangzhou Diabetic Eye Study and healthy controls from the community of Guangzhou, China were recruited in this cross-sectional study. Demographic, clinical and ocular parameters were regularly obtained. Retinal thickness (RT), retinal nerve fiber layer (RNFL) thickness, ganglion cell-inner plexiform layer (GC-IPL) thickness, outer retina layer (ORL) thickness and choroid thickness (CT) were automatically evaluated by swept-source optical coherence tomography (SS-OCT) in the 1, 3, and 6 mm centered on fovea. Vessel density (VD) was obtained by OCT angiography (OCTA) in the 1 and 3 mm centered on the fovea.

**Results:** 1,444 eyes of 1,444 individuals (722 T2DM patients and 722 healthy controls) were included in the final analyses. Macular average RNFL was thinned (P < 0.001), average GC-IPL was thickened (P < 0.001), and ORL was thickened (P = 0.012) in DM patients compared with healthy controls independent of confounding factors. VD was significantly increased in diabetic patients without DR. Correlations were found between VD and specific layers of retina both in DM patients and in healthy controls. Especially in DM patients, average RT in positively related with parafoveal VD (β= 0.010, 95%CI: 0.003 to 0.017) and total average VD (β= 0.010,95%CI: 0.003 to 0.016). Specifically, RNFL thickness is inversely related with VD, while both GC-IPL and ORL thickness are positively related with VD in diabetic patients without DR.

**Conclusion:** This study reports and correlates the early alterations of chorioretinal structure and retinal superficial vessels in T2DM patients even before the onset of clinical signs of DR. Findings of this study may provide novel insights to explore the pathogenesis of DR.

## Introduction

Diabetic retinopathy (DR) is one of the most common complications of diabetic mellitus (DM).^1^ By 2040, there will be 93 million DR patients, 38 million of which may occur vision threatening DR.^2^ However, the pathogenesis of DR is still not fully demonstrated. Systemic factors (e.g. the course of DM, poor control of blood sugar, hypertension, and hyperlipidemia) were reported to relate with the onset of DR, but these factors can only explain less than 10% of the onset risk. ^3, 4^ A noteworthy phenomenon is that some DM patients do not suffer from DR by long time, while others suffer from vision threatened DR in a short period of time, i.e. dramatic variability exists in development and progression of DR among different subjects, which is not satisfied interpreted through the aforementioned factors, neither. Therefore, identifying the difference between healthy populations and DM patients without DR may attach great importance to understand the mechanism behind the pathogenesis of DR.

DR is featured as changes of the neurovascular unit (NVU) of the retina.^5^ Chronic hyperglycemia may cause oxidative stress, hypoxia, and inflammation, which leads to the disruption of NVU, and ends up as morphological chorioretinal alterations in DR patients.^6^ The introduction of optical coherence tomography (OCT) and OCT angiography (OCTA) provides us with a non-invasive method to observe not only anatomical features but also functional alterations of the chorioretinal in DM patients.^7^Several studies assessed the morphological alteration of chorioretinal structures in DM patients with and without DR.^8-20^ Previous studies also reported that the alterations of blood flow occurred earlier than distinctive pathologic changes of DR (e.g. the micro aneurysm).^19-23^ However, the results above remained controversial due to varied reasons including the limitation of techniques, sample size, and confounding factors. Based on the observed alterations, Cunha-Vas et al raised the definition of three pathologic phenotypes in the initial stage of DR, which are characterized by three main pathways of the disease: neurodegeneration, edema, and ischemia.^24, 25^ Neurodegeneration can be recognized as the thinning of retinal nerve fiber layer (RNFL) and ganglion cell-inner plexiform layer (GC-IPL); edema can be recognized as the thickening of inner nuclear layer, outer plexiform layer or the full retina layer; ischemia can be recognized as the alteration of the vessel density.^24^

It is traditionally supposed that DR is a microvascular disease, while more and more studies reported that neuropathy existed with the absence of vasculopathy.^26, 27^ It suggested that the dysfunction of neurovascular interaction may be a key factor before the onset of clinically evident DR.^5^ However, the data of early evaluation of the correlation between microvascular changes detected on OCTA and chorioretinal layer thickness changes detected on OCT in DM patients with no DR signs is very limited.^15, 28, 29^ Furthermore, the results of aforementioned studies also needs to be verified by more data due to the limitations of relatively small sample size and the inadequate inclusion of potential confounding factors. Therefore, the aim of this study was to evaluate and correlate the alterations of chorioretinal layer thickness detected on OCT and early microvascular changes detected on OCTA in a relatively large scale of patients with Type 2 DM without clinical signs of DR.

## Methods

### Participants

This is a cross-sectional study performed at the Zhongshan Ophthalmic Centre (ZOC), Sun Yat-sen University, Guangzhou, China. The Institute Ethics Committee of ZOC approved the protocol of this study. The study was conducted according to the tenets of the Helsinki Declaration. Written informed consent was obtained to each participants before attending the study. Healthy participants (NDM group) were recruited from the community in Guangzhou. The inclusion criteria of normal controls are: (1) have no history of systemic diseases or operations; (2) have no history of ocular diseases or operations; (3) have normal results of blood tests; (4) have normal results of ocular examination in both anterior and posterior segments; (5) axial length (AL) is less than 26 mm, i.e. have no high myopia. DM patients (DM group) were randomized sampled from the Guangzhou Diabetic Eye Study (GDES) cohort, which is a community-based prospective cohort study that includes type 2 diabetic patients registered in the community health centres near ZOC. Type 2 DM (T2DM) patients met the following criteria were randomized in this study: (1) have no history of systemic diseases or operations; (2) have no history of ocular diseases or operations; (3) have no clinical signs of DR.

### Systemic and laboratory parameters

Demographic information was obtained by questionnaires, including age, sex, and course of diabetes. Height, weight and blood pressure (BP) were measured by an experienced nurse. Body mass index (BMI) = weight(kg)/height^2^ (m^2^). Mean BP = Diastolic BP + 1/3 (Systolic BP – Diastolic BP). Laboratory parameters, including hemoglobin A1c (HbA1c), triglycerides (TG), total cholesterol (TC), low-density lipoproteins (LDL), high-density lipoproteins (HDL), serum uric acid (UA), serum creatinine (SCr), and C-reactive protein (CRP) were examined in a standardized laboratory.

### Ocular examination

All participants received comprehensive ocular examinations. The anterior segments were evaluated by slit-lamp biomicroscopy. The posterior segments were evaluated by ophthalmoscopy. Best corrected visual acuity (BCVA) were measure with the Early Treatment of Diabetic Retinopathy Study (ETDRS) LogMAR E chart (Precision Vision, Villa Park, Illinois, USA). Refraction were measured after mydriasis with an auto refractometer (KR-8800; Topcon, Japan). Intraocular pressure (IOP) was measured after mydriasis using Topcon CT-80A non-contact tonometry (Topcon, Japan). Ocular biometric parameters were measured using Lenstar LS900 (HAAG-Streit AG, Koeniz, Switzerland) in dark conditions.

### SS-OCT imaging

The retina and choroid was examined with SS-OCT (DRI OCT-2 Triton; Topcon, Tokyo, Japan) to perform cross-sectional scanning of the macular retina and choroid. A 6*6 mm volume scan protocol, which presents the thicknesses of each layer of retina and choroid in 9-ETDRS sectors, was used to scan the macular region. Segmentation of different layers were automatically performed by the built-in software program. The result of segmentation was inspected and confirmed by one OCT technician, performing manual correction if the software misjudged the borderline of each layer. Parameters assessed by SS-OCT are as following. Retinal nerve fiber layer (RNFL) thickness was defined as the distance from inner limiting membrane to RNFL. Ganglion cell-inner plexiform layer (GC-IPL) thickness was defined as the distance from the borderline between the RNFL and ganglion cell layer to the borderline between the inner plexiform layer and inner nuclear layer. Outer retinal layer (ORL) thickness was defined as the distance from the borderline between the inner plexiform layer and inner nuclear layer to the borderline between the retinal pigment epithelium and the Bruch membrane. Choroidal thickness (CT) and retinal thickness (RT) were defined as usual. Macular OCT image were evaluated as three regions: foveal (thickness of circular area with 1 mm diameter centered on the fovea), inner ring (mean thickness of 4 inner quadrants with diameter of 3 mm), and outer ring (mean thickness of 4 outer quadrants with diameter of 6 mm). (Figure 1A & B)

**Figure 1.**
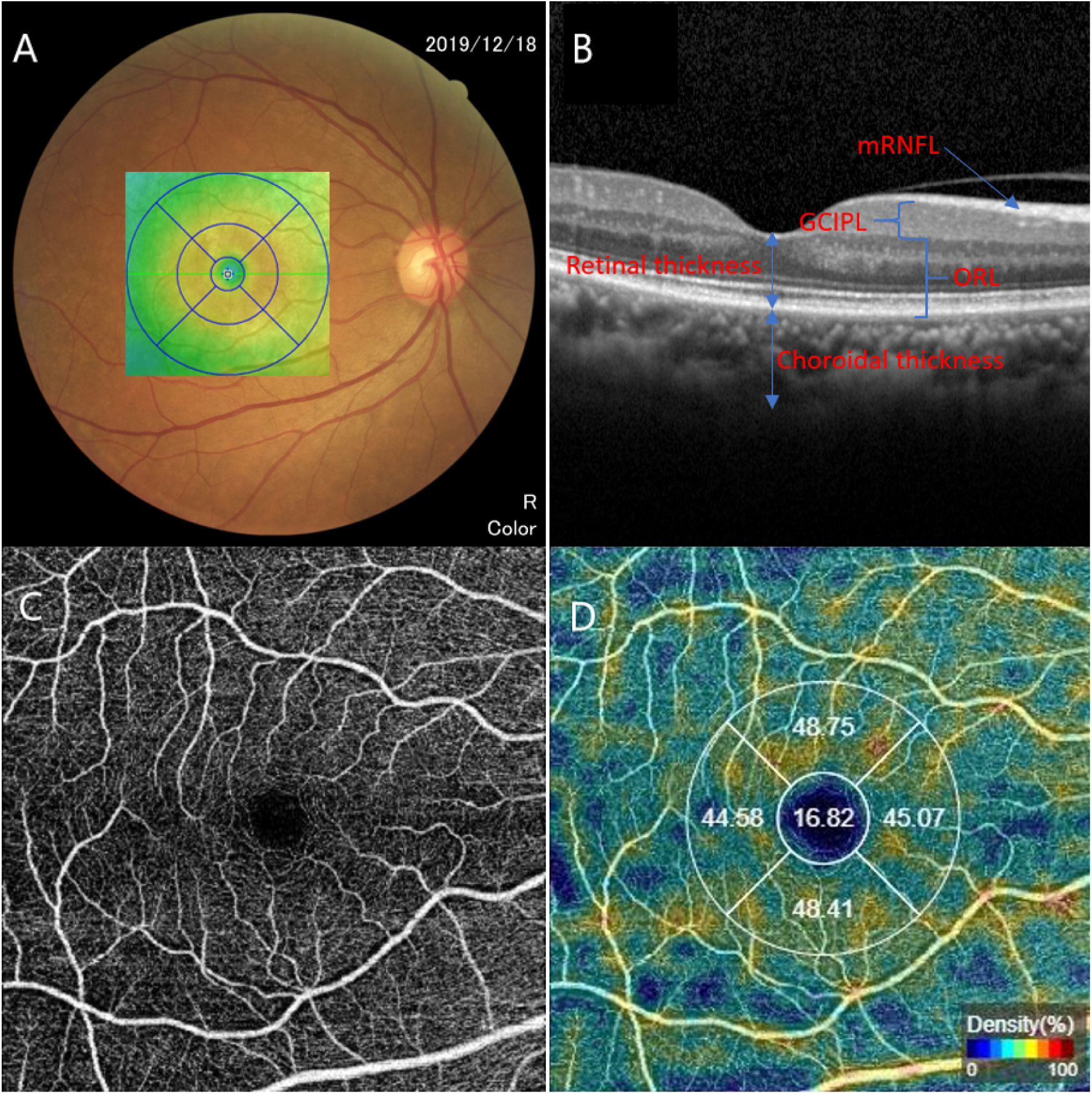
Illustrations of the OCT and OCTA measurements. VD = vessel density; mRT = macular retinal thickness; mRNFL = macular retinal nerve fiber layer; GC-IPL = ganglion cell-inner plexiform layer; ORL = outer retinal layer; mCT = macular choroid thickness.

### OCTA evaluation

Blood flow of the superficial layer of retina was obtained by a swept source OCTA device (Triton DRI-OCT, Topcon. Inc., Tokyo, Japan), which provided a wavelength of 1050 nm within a range of 100 nm and an achievable horizontal resolution of 8 μm. Blood flow was visualized using the 3×3mm model. The vessel density (VD) of the superficial layer was also automatically calculated by the built-in software. Parafoveal average VD was defined as the mean VD of 4 quadrants (temporal, superior, nasal, and inferior) with diameter of 3 mm. (Figure 1C & D)

### Statistical analyses

Data of right eye from each participant were incorporated into the final analyses. Normal distribution was firstly verified using the Kolmogorov-Smirnov test. Inter-group differences of demographic, systemic and ocular quantitative parameters were evaluated by t-test. The chi-square test was used for identify the difference of sex proportion between the two groups. Then, we performed multivariable regression to adjust the effect caused by potential confounding factors. We adjusted age and sex in model 1. We further adjusted age, sex, BMI, mean BP, TC, HbA1c, TG, SCr, and AL in model 2. The correlations of VD with OCT parameters were analysed using linear regression. The confounding factors adjusted in multivariable linear regression were the same as those in model 2. A P value of < 0.05 was considered to be significant. All the above analyses were conducted using Stata Version 14.0 (Stata Corporation, College Station, TX, USA).

## Results

### Demographic and clinical features

Of the 1,444 participants included in the final statistical analysis, half of them (n = 722) were healthy subjects (NDM group) and the other half of them (n = 722) were T2DM patients without clinical signs of diabetic retinopathy (DM group). Table 1 reports the basic demographic and clinical characteristics of the participants. Among all the participants, 980 (67.87%) of them were female, the average age was 57.99 ± 7.57 years. 722 DM patients were more likely to be male (P < 0.001), and were older (P < 0.001) compared to the 722 healthy controls. The mean course of diabetes in DM group was 6.85±5.58 years. No differences were found in SE, AL, corneal curvature, and corneal diameter between the groups (all, P > 0.05). Other parameters were significantly different between the groups, detailed in Table 1.

**Table 1.** Demographic and clinical characteristics of the participants.

| Characteristics | Overall | NDM | DM | P |
| --- | --- | --- | --- | --- |
| No. of participants | 1444 | 722 | 722 | - |
| Female, % | 980(67.87) | 546(75.62) | 434(60.11) | **<0.001** |
| Mean age, year | 57.99±7.57 | 57.07±9.14 | 58.90±5.43 | **<0.001** |
| Duration of diabetes, year | - | - | 6.85±5.58 | - |
| Body mass index, kg/m<sup>2</sup> | 24.01±3.40 | 23.28±3.05 | 24.73±3.57 | **<0.001** |
| Mean BP, mm Hg | 175.14±24.05 | 170.23±24.11 | 180.05±22.97 | **<0.001** |
| Hemoglobin A1c, % | 6.38±1.25 | 5.76±0.58 | 6.99±1.42 | **<0.001** |
| Triglycerides, mmol/L | 2.28±1.68 | 2.11±1.53 | 2.46±1.80 | **<0.001** |
| Total cholesterol, mmol/L | 5.08±1.01 | 5.21±0.99 | 4.94±1.01 | **<0.001** |
| Cholesterol/triglycerides ratio | 3.11±1.76 | 3.43±1.86 | 2.80±1.60 | **<0.001** |
| Low-density lipoproteins, mmol/L | 3.20±0.88 | 3.27±0.84 | 3.13±0.92 | **0.002** |
| High-density lipoproteins, mmol/L | 1.33±0.42 | 1.38±0.43 | 1.28±0.41 | **<0.001** |
| Serum uric acid, µmol/L | 362.86±94.82 | 349.37±86.89 | 376.37±100.41 | **<0.001** |
| Serum creatinine, µmol/L | 67.73±16.32 | 67.58±14.59 | 67.88±17.89 | 0.728 |
| C-reactive protein, mg/L | 2.20±4.81 | 1.71±2.65 | 2.68±6.23 | **<0.001** |
| BCVA, logMAR | 82.91±3.82 | 83.06±3.84 | 82.41±3.72 | **0.028** |
| Spherical equivalent, D | 0.16±1.93 | 0.20±1.97 | 0.13±1.89 | 0.511 |
| Intraocular pressure, mm Hg | 16.20±2.53 | 15.72±2.29 | 16.68±2.66 | **<0.001** |
| Lens thickness, mm | 4.57±0.34 | 4.52±0.35 | 4.63±0.33 | **<0.001** |
| Axial length, mm | 23.44±0.92 | 23.45±0.92 | 23.44±0.93 | 0.757 |
| Corneal curvature, D | 44.15±1.40 | 44.12±1.34 | 44.18±1.45 | 0.377 |
| Corneal diameter, mm | 11.67±0.39 | 11.69±0.38 | 11.65±0.39 | 0.096 |

Data are expressed as the mean ± SD.
NDM = participants without diabetes mellitus; DM = diabetic participants without diabetic retinopathy; BP = blood pressure; Mean BP = Diastolic BP + 1/3 (Systolic BP – Diastolic BP)
P-value less than 0.05 were shown in bold.

### Retina and microcirculation alterations

The chorioretinal thickness of different layers in various regions was shown in Table 2. The average RT did not differ between the groups. The average RNFL thickness of diabetic eyes was significantly thinner than NDM subjects (P < 0.001). The average thickness of GC-IPL was thicker in DM patients (73.02±6.41 μm vs 71.48±5.02 μm, P < 0.001). As to the ORL in DM patients, average thickness was thicker (171.94±32.16 μm vs 168.26±8.58 μm, P = 0.004). Moreover, thickness of macular choroid only differed in the region of outer ring, which was thinner in diabetic patients (P = 0.012). After adjusting for age and sex in model 1, diabetic patients also had chorioretinal alterations compared with NDM participants, as shown in Table 3. Retina was thicker in foveal (P = 0.041), and significantly thinner in outer ring and inner ring (both, P < 0.001). Average RNFL remained thinner (P < 0.001). Average and foveal thickness of GC-IPL were thicker (P < 0.001 and P = 0.007, respectively). ORL was thicker in average (P = 0.012), and significantly thinner in outer ring and inner ring (both, P < 0.001). Macular choroid was only thinned in outer ring (P = 0.039). We further adjusted other confounding variables in model 2 (including age, sex, BMI, mean BP, TC, HbA1c, TG, SCr, and AL), showing that most of the alterations still remained statistically significant, as it shown in Table 3.

**Table 2.** OCT and OCTA parameters between the participants without and with DM.

| parameters | Overall | NDM | DM | P |
| --- | --- | --- | --- | --- |
| **Macular Retinal thickness, $\mu\text{m}$** |  |  |  |  |
| Average | 275.40±23.44 | 274.92±13.08 | 276.11±33.28 | 0.385 |
| Foveal | 227.45±27.23 | 225.55±21.60 | 230.27±33.74 | **0.003** |
| Outer ring | 248.72±66.99 | 268.40±13.47 | 225.03±93.01 | **<0.001** |
| Inner ring | 278.46±80.40 | 303.06±15.37 | 248.84±111.20 | **<0.001** |
| **Macular RNFL thickness, $\mu\text{m}$** |  |  |  |  |
| Average | 33.48±4.57 | 35.20±3.49 | 31.43±4.86 | **<0.001** |
| Foveal | 23.94±8.66 | 20.20±2.74 | 24.88±9.45 | 0.423 |
| Outer ring | 39.51±5.41 | 39.40±4.20 | 39.63±6.57 | 0.441 |
| Inner ring | 25.02±2.74 | 24.95±2.55 | 25.11±2.95 | 0.305 |
| **GC-IPL thickness, $\mu\text{m}$** |  |  |  |  |
| Average | 72.18±5.74 | 71.48±5.02 | 73.02±6.41 | **<0.001** |
| Foveal | 39.99±8.77 | 39.16±7.76 | 40.97±9.75 | **<0.001** |
| Outer ring | 67.51±5.39 | 67.44±5.13 | 67.59±5.70 | 0.603 |
| Inner ring | 88.95±7.71 | 89.16±7.13 | 88.71±8.35 | 0.290 |
| **Outer retinal layer thickness, $\mu\text{m}$** |  |  |  |  |
| Average | 169.75±21.54 | 168.26±8.58 | 171.94±32.16 | **0.004** |
| Foveal | 184.59±22.35 | 184.09±14.39 | 185.33±30.59 | 0.345 |
| Outer ring | 141.74±66.05 | 161.55±8.75 | 117.86±92.17 | **<0.001** |
| Inner ring | 164.44±79.45 | 188.95±9.73 | 134.90±110.55 | **<0.001** |
| **Macular choroidal thickness, $\mu\text{m}$** |  |  |  |  |
| Average | 210.41±69.49 | 212.13±68.72 | 208.36±70.40 | 0.326 |
| Foveal | 231.10±82.01 | 233.68±81.74 | 228.04±82.30 | 0.213 |
| Outer ring | 202.60±67.57 | 206.89±67.24 | 197.52±67.66 | **0.012** |
| Inner ring | 224.11±75.81 | 227.09±75.86 | 220.58±75.66 | 0.120 |
| **Vessel density, %** |  |  |  |  |
| Parafoveal average | 46.46±2.77 | 45.42±2.28 | 47.53±2.82 | **<0.001** |
| Total | 43.33±2.54 | 42.31±2.05 | 44.36±2.57 | **<0.001** |
| Temporal | 46.61±3.69 | 45.25±2.67 | 47.98±4.06 | **<0.001** |
| Superior | 48.16±3.89 | 47.26±3.36 | 49.07±4.17 | **<0.001** |
| Nasal | 44.23±3.53 | 43.51±3.48 | 44.97±3.43 | **<0.001** |
| Inferior | 46.86±4.24 | 45.64±3.59 | 48.10±4.48 | **<0.001** |

Data are expressed as the mean ± SD.
NDM = participants without diabetes mellitus; DM = diabetic participants without diabetic retinopathy; RNFL = retinal nerve fiber layer; GC-IPL = Ganglion Cell-Inner Plexiform Layer.
P-value less than 0.05 were shown in bold.

**Table 3.** Differences of OCT and OCTA parameters between the participants without and with DM after adjusting confounding variables.

| NDM vs DM | Model 1<sup>*</sup> |  | Model 2<sup>†</sup> |  |
| --- | --- | --- | --- | --- |
|  | β (95% CI) | P | β (95% CI) | P |
| **Macular Retinal thickness, μm** |  |  |  |  |
| Average | 0.98(-1.74, 3.69) | 0.480 | 1.13(-2.15, 4.42) | 0.498 |
| Foveal | 3.28(0.13, 6.43) | **0.041** | 3.90(0.09, 7.72) | **0.045** |
| Outer ring | -43.43(-50.40, -36.45) | **<0.001** | -48.48(-56.81, -40.16) | **<0.001** |
| Inner ring | -55.11(-63.43, -46.78) | **<0.001** | -60.65(-70.58, -50.71) | **<0.001** |
| **Macular RNFL thickness, μm** |  |  |  |  |
| Average | -3.55(-4.00, -3.09) | **<0.001** | -3.49(-4.03, -2.95) | **<0.001** |
| Foveal | 5.19(-10.36, 20.74) | 0.478 | 19.13(-14.70, 52.97) | 0.192 |
| Outer ring | 0.56(-0.31, 1.15) | 0.063 | 0.58(-0.12, 1.28) | 0.106 |
| Inner ring | 0.12(-0.18, 0.42) | 0.421 | 0.10(-0.25, 0.46) | 0.559 |
| **GC-IPL thickness, μm** |  |  |  |  |
| Average | 1.65(1.03, 2.27) | **<0.001** | 1.67(0.93, 2.40) | **<0.001** |
| Foveal | 1.30(0.36, 2.24) | **0.007** | 1.05(-0.07, 2.16) | 0.066 |
| Outer ring | 0.40(-0.19, 0.99) | 0.179 | 0.40(-0.26, 1.06) | 0.232 |
| Inner ring | -0.43(-1.27, 0.41) | 0.314 | -0.26(-1.07, 0.74) | 0.605 |
| **Outer retinal layer thickness, μm** |  |  |  |  |
| Average | 3.20(0.71, 5.71) | **0.012** | 3.45(0.41, 6.49) | **0.026** |
| Foveal | 0.52(-2.10, 3.14) | 0.698 | 1.49(-1.69, 4.68) | 0.358 |
| Outer ring | -44.27(-51.14, -37.41) | **<0.001** | -49.34(-57.54, -41.14) | **<0.001** |
| Inner ring | -54.88(-63.11, -46.64) | **<0.001** | -60.48(-70.31, -50.65) | **<0.001** |
| **Macular choroidal thickness, μm** |  |  |  |  |
| Average | -2.40(-9.80, 5.00) | 0.525 | -2.46(-10.80, 5.87) | 0.562 |
| Foveal | -6.28(-15.14, 2.58) | 0.164 | -7.05(-17.10, 3.00) | 0.169 |
| Outer ring | -7.54(-14.69, -0.40) | **0.039** | -6.75(-14.82, 1.31) | 0.101 |
| Inner ring | -5.95(-14.10, 2.21) | 0.153 | -6.73(-15.96, 2.50) | 0.153 |
| **Vessel density, %** |  |  |  |  |
| Parafoveal average | 2.24(1.96, 2.52) | **<0.001** | 2.31(1.98, 2.64) | **<0.001** |
| Total | 2.16(1.91, 2.41) | **<0.001** | 2.17(1.87, 2.46) | **<0.001** |
| Temporal | 2.85(2.47, 3.22) | **<0.001** | 2.84(2.39, 3.29) | **<0.001** |
| Superior | 1.99(1.58, 2.41) | **<0.001** | 2.30(1.81, 2.79) | **<0.001** |
| Nasal | 1.61(1.24, 1.99) | **<0.001** | 1.64(1.20, 2.09) | **<0.001** |
| Inferior | 2.49(2.05, 2.94) | **<0.001** | 2.46(1.82, 2.99) | **<0.001** |

\*Model 1 adjusted for age and sex.
†Model 2 adjusted for age, sex, body mass index, mean BP, total cholesterol, hemoglobin A1c, triglycerides, serum creatinine, and axial length.
P-value less than 0.05 were shown in bold.

Microcirculation status was assessed based on the VD data of OCTA, results were reported in Table 2. In particular, VD was found significantly higher in diabetic patients compared with healthy controls, no matter the average data or in each subfields (all, P < 0.001). After adjusting for the potential confounding factors, the vessel density remained significantly higher in DM patients (all, P < 0.001), as it presented in Table 3.

### Association between chorioretinal thickness and VD

Table 4 presents the associations of average chorioretinal thickness of different layers with VD. Most of the OCT parameters were correlated with VD. It persistently revealed that macular RT and VD had positive association independent of confounding factors, with the exception of no correlation of average macular RT with parafoveal VD in NDM group. Macular RNFL thickness was negatively correlated with VD in all participants as well as in DM patients, which is persistently significant after adjusting for confounding factors. However, the positive correlation between RNFL thickness and VD is positive in healthy subjects is no longer significant in the multivariable model. As to the GC-IPL, it persistently revealed that average GC-IPL thickness is positively associated with VD in all participants independent of confounding factors. As to the ORL thickness, it is positively correlated with VD in the whole participants and in DM patients, but showed no association in healthy subjects. The average macular CT showed weak correlation with VD, which is only significant in healthy subjects when confounding factors were taken into analysis.

**Table 4.** Univariable and multivariable regression analysis of association between OCTA parameters and OCT parameters.

| $\beta$ (95% CI) | average mRT | average mRNFL thickness | average GC-IPL thickness | average ORL thickness | average mCT |
| --- | --- | --- | --- | --- | --- |
| **Univariable regression** |  |  |  |  |  |
| **Overall** |  |  |  |  |  |
| parafoveal VD | 0.013(0.006, 0.019) | -0.163(-0.195, -0.132) | 0.103(0.078, 0.129) | 0.015(0.008, 0.022) | 0.002(0.000,0.005) |
| total average VD | 0.130(0.007, 0.019) | -0.149(-0.178, -0.120) | 0.105(0.081, 0.128) | 0.015(0.008, 0.021) | 0.002(-0.000,0.004) |
| **NDM** |  |  |  |  |  |
| parafoveal VD | 0.019(0.006, 0.033) | 0.057(0.008, 0.107) | 0.050(0.015, 0.085) | 0.017(-0.003, 0.037) | 0.006(0.003, 0.008) |
| total average VD | 0.021(0.009, 0.032) | 0.071(0.027, 0.115) | 0.057(0.025, 0.880) | 0.016(-0.002, 0.034) | 0.005(0.002, 0.007) |
| **DM** |  |  |  |  |  |
| parafoveal VD | 0.010(0.003, 0.017) | -0.177(-0.220, -0.133) | 0.104(0.070, 0.138) | 0.010(0.003, 0.018) | -0.001(-0.004, -0.003) |
| total average VD | 0.010(0.004, 0.017) | -0.161(-0.202, -0.121) | 0.103(0.072, 0.134) | 0.010(0.003, 0.017) | -0.001(-0.004, -0.002) |
| **Multivariable regression<sup>†</sup>** |  |  |  |  |  |
| **Overall** |  |  |  |  |  |
| parafoveal VD | 0.010(0.004, 0.017) | -0.170(-0.203, -0.138) | 0.103(0.077, 0.129) | 0.012(0.005, 0.019) | 0.002(-0.000, 0.004) |
| total average VD | 0.011(0.005, 0.017) | -0.155(-0.184, -0.125) | 0.105(0.081, 0.128) | 0.012(0.005, 0.018) | 0.001(-0.001, 0.004) |
| **NDM** |  |  |  |  |  |
| parafoveal VD | 0.010(-0.004, 0.024) | 0.019(-0.030, 0.069) | **0.047(0.011, 0.083)** | 0.004(-0.017, 0.025) | **0.005(0.002, 0.007)** |
| total average VD | **0.013(0.000, 0.025)** | 0.034(-0.011, 0.079) | **0.056(0.024, 0.088)** | 0.004(-0.015, 0.023) | **0.004(0.001, 0.006)** |
| **DM** |  |  |  |  |  |
| parafoveal VD | 0.010(0.003, 0.017) | -0.187(-0.231, -0.142) | 0.105(0.070, 0.140) | 0.010(0.003, 0.017) | -0.000(-0.004, 0.003) |
| total average VD | 0.010(0.003, 0.016) | -0.173(-0.214, -0.132) | 0.104(0.073, 0.136) | 0.010(0.003, 0.016) | -0.001(-0.004, 0.003) |

Bold indicates statistically significant ( $P < 0.05$ ).
VD = vessel density; mRT = macular retinal thickness; mRNFL = macular retinal nerve fiber layer; GC-IPL = ganglion cell-inner plexiform layer; ORL = outer retinal layer; mCT = macular choroid thickness.
†Multivariable regression model adjusted for age, sex, body mass index, mean BP, total cholesterol, hemoglobin A1c, triglycerides, serum creatinine, and axial length.

## Discussion

The present study explored the alterations of chorioretinal layer thickness detected on OCT and early microvascular changes detected on OCTA in a relatively large scale of patients with Type 2 DM without clinical signs of DR. Our results suggested that the alterations of chorioretinal thickness and retinal vessel density could be detected by OCT and OCTA before the onset of clinical signs of DR in T2DM patients. In particular, macular RNFL was significantly thinned, GC-IPL thickened, ORL thickened, VD increased in DM patients compared with healthy controls. Furthermore, there were correlations between VD and specific layers of retina both in DM patients and in healthy controls. Especially in DM patients, average RT is positively related with VD. Specifically, RNFL thickness is inversely related with VD, and both GC-IPL and ORL thickness are positively related with VD. To the best of our knowledge, this is the first study to evaluate the status and the association of chorioretinal layer thickness and early microvascular changes in Chinese community population of T2DM patients without DR.

DR is a disease of neurovascular unit of retina.^6^ Publications also reported that chronic hyperglycemia in DM may alter the component of neurovascular even before the clinical signs of DR.^30-34^ Therefore, it is reasonable to raise the hypothesis that there may be morphological changes in retinal structures. Our findings suggest that, during the initial period of retina neurovascular alteration of DM, retinal nerve fiber atrophied, while the structure from RPE and ganglion cell were all hydropic, accompanying the increased blood flow of the superficial retina. Together with the correlation of VD with retinal thickness we found in this study, we speculate that hyperglycemia may firstly induce the atrophy of retinal nerve fiber and cellular swelling of the retinal cell bodies, which may furtherly cause compensatory increasing of retinal superficial blood flow. However, considering the cross-sectional design of this study, longitudinal studies are warranted to verify the speculation we raised.

Changes of chorioretinal layers in the macula in DM without DR or with the early stage of DR has been reported in the previous publications. Several studies reported a significantly thinner RNFL thickness in T2DM^8-10, 12, 14, 16, 28, 35^, which is further confirmed in our study. These results suggested that neurodegeneration occurred in the initial stage of DR, which may be an early event in the pathogenesis of DR shortly after the disruption of the NVU.

Alterations of ganglion cells also represents neurodegeneration. Vujosevic et al^36^ reported that the thickness from inner limiting membrane/RNFL interface to inner plexiform layer/inner nuclear layer interface (i.e. the RNFL and GC-IPL) in central subfoveal differed among 3 examined groups (healthy controls, T1DM, and T2DM). T1DM patients were likely to be thicker, while T2DM patients had the potential to be thinner compared to the controls independent of age, though were not statistically significant. Interestingly, we found GC-IPL thickness significantly increased in foveal region in T2DM, independent of age and sex. However, it is not significant any more after adjusting for further factors. Pierro et al^13^ documented thinned GCL thickness in T2DM patients without DR. Van Dijk et al^10^ identified decreased GCL and IPL thickness only in patients with minimal DR compared with controls. However, we found increased GC-IPL thickness in T2DM patients, which is inconsistent with the aforementioned studies. It is believed that ganglion cells and amacrine cells are firstly influenced by diabetes, which is formerly detected to be firstly apoptosis.^5^ However, our results did not support this point of view. Considering our results of thinning RNFL, we speculated that the damage of NVU might firstly affect the retinal nerve fiber, and ganglion cell body may be dropsy at the same time. Though the thinning of GC-IPL indicates neurodegeneration^24^, our findings suggested that ganglion cell edema may be an earlier cellular pathological process before it apoptosis.

Outer retinal layer (ORL), including inner nuclear layer (INL), outer plexiform layer (OPL), outer nuclear layer (ONL), ellipsoid zone (EZ), and retinal pigment epithelium RPE, was reported to be a biomarker for retinal diseases including retinitis pigmentosa, Stargardt disease, and retinopathy of prematurity.^37, 38^ Recent studies in animal models suggested that microvascular damage may affect retinal photoreceptor cells, which may be a crucial role in the pathogenesis of DM.^39^ It also reported that in experimental models, photoreceptors also have an increased apoptotic rate besides the ganglion cells and amacrine cells.^5^ There are limited studies in vivo to assess the ORL. Sacconi et al reported no significant changes in foveal OPL, ONL, and EZ in T1DM without funduscopic signs of DR.^23^ Data in our study also indicated no significant alterations in foveal ORL in T2DM patients. However, it is worthwhile to note that obvious decreased ORL thickness was found in the both outer and inner rings and average ORL thickness was found to be significantly increased in this study, which indicates that the photoreceptor may change in vivo in T2DM patients. Though the exact mechanism of different situations of photoreceptor in different regions is not to be fully interpreted by us, we speculated that hyperglycemia may affect rod cells more due to the uneven distribution of photoreceptors between the foveal and the subfoveal regions.

As to the choroid, Esmaeelpour et al,^40^ Vujosevic et al,^41^ and Querques et al^42^ reported choroidal alterations in diabetic patients without DR, Tavares Ferreira et al^43^ reported increased but not statistically significant CT in diabetic patients without DR. However, we found no significant alterations of macular CT. Together with the changes of ORL we found, we speculated that the outer layers of retina, mainly supplied by choroid, may have morphological changes even before CT changes. It suggested that the alteration of ORL and CT before the onset of clinical signs of DR is a metabolic process. Thus, the interaction of choroid and outer layers of retina in early stage of DR pathogenesis is still under debate, which is warranted more studies to demonstrate. Even so, based on existing findings, it is necessary to have early OCT examinations for T2DM patients in order to early prevent, diagnose, and treat DR. Blood flow is critical in DR pathogenesis. How does blood flow affect DR in still controversial, which is reported to be decreased or to be increased in diabetic retinal vessels.^15^ Changes of macular capillary network have been reported in DM without DR.^15, 21, 28, 29, 44^ De Carlos et al^44^ identified more prevalent alterations to the foveal avascular zone and capillary non-perfusion in diabetic eyes. Dimitrova et al^15^ identified decreasing superficial and deep retinal VD in parafovea region in diabetic patients without DR compared to healthy subjects. On the contrary, Rosen et al^21^ reported increased perfused capillary density values in the No DR group compared with healthy controls. We also found a higher VD in the parafovea region in diabetic eyes without DR independent of confounding factors, which is considered to be an autoregulatory response to increased metabolic demand, i.e. a consequence of compensation. This may provide a novel angel to explain the pathogenesis of DR.

As a retinal NVU disease, the coupling dysfunction of neurovascular components may be a critical factor in the progression of DR.^5^ Thus, investigating the association between VD and retinal structure in the early stage of DR is meaningful. Vujosevic et al^28^ documented that perifoveal capillary loss in superficial capillary and inner retinal layer thickness had significant correlation in both T1DM and T2DM. Dimitrova et al^15^ reported that superficial vessel density significantly correlated with full RT in parafovea of healthy subjects, but had no significant association in diabetic eyes. However, the previous two studies only considered limited confounding factors. We adjusted more confounding factors (including BMI, TC, TG, SCr, HbA1c, etc.) when assessing the correlation, finding that VD was positively correlated with alterations of all OCT parameters, which suggested that vasculopathy plays an essential role in changes of different retinal layers. Specifically, VD was positively correlated with full RT, GC-IPL thickness and ORL thickness, and it was negatively correlated with RNFL thickness. But the causal relationship between the microcirculation and different retina layers need to be confirmed by further longitudinal studies. And the exact mechanism behind these findings is still unclear. Blood-retinal barrier (BRB) dysfunction may be a potential mechanism. Considering the different correlation in different layers, we speculated that BRB dysfunction may affect different layers via independent pathways, which are warranted to be investigated through further fundamental studies.

This study surely have certain limitations. First, because of the nature of this cross-sectional study, it is not supposed to determine whether the neural alterations or the vascular alterations preceded. The sequential order of the alterations of different chorioretinal layers is also difficult to determine, too. Secondly, we only obtained the superficial VD in this research. Nevertheless, the microcirculation of deep capillary plexus may also cause chorioretinal structure changes, which would be clarified in our further studies aimed on this specific aspect. Thirdly, we only included T2DM subjects, but the alterations of retinal neurovascular structure are quite diverse in T1DM and T2DM.^15, 28, 29, 44^ Thus, the conclusions of our study were only applicable to T2DM patients, which is not appropriate to extend to T1DM patients. Finally, the basic characteristics of the two groups are not balanced enough. DM patients were more likely to be male, and were older. Moreover, most parameters we collected had statistical differences between the groups. Considering the reported factors that may affect chorioretinal thickness and VD, we adequately adjusted for age, sex, body mass index, mean BP, total cholesterol, hemoglobin A1c, triglycerides, serum creatinine, and axial length in the multivariable models, which made our results more credible.

## Conclusion

In summary, this study reports and correlates the early alterations of chorioretinal structure and retinal superficial vessels in T2DM patients before the onset of clinical signs of DR. Findings of this study may provide novel insights to explore the pathogenesis of DR. Early OCT and OCTA detection are necessary for DM patients without DR. Longitudinal and larger scale studies are needed to further explain the changes and correlations of neurovascular changes in the diabetic retina.

## Data Availability

The availability of all data referred to in the manuscript and note links below.

